# Integrating Wearable Sensor Technology into Outpatient Rehabilitation Care: A Proof-of-Concept Study

**DOI:** 10.64898/2026.09.04.26362284

**Authors:** Allison E. Miller, Carey L. Holleran, Marghuretta D. Bland, Mary M. Crumley, Brandon J. Jensen, Keith R. Lohse, Ellen E. Fitzsimmons-Craft, Caitlin A. Newman, Thomas M. Maddox, Catherine E. Lang

## Abstract

**Background:** The disconnect between episodic in-clinic assessments and patients’ goals to improve activity in daily life presents a challenge in rehabilitation. Wearable sensors are promising for addressing this disconnect but are not part of routine practice. The purpose of this study was to develop and test a digital health solution that integrated wearable sensor technology into outpatient rehabilitation.

**Methods:** Using an iterative user-centered design approach, a digital solution was built that integrated data from a wrist sensor into the electronic health record (EHR). The feasibility, usability, and resources required of the solution were evaluated in a 9-month proof-of-concept study in outpatient rehabilitation.

**Results:** 17 clinicians and 38 patients (median monitoring duration: 42 days) participated. Patient adherence to daily data syncing was a median of 68.6% and was not significantly correlated with digital technology self-efficacy (*r* = 0.03, *p* = 0.86). Recent sensor data (≤ 3 days old) were available in the EHR for 90.2% (193/214) of clinic visits, and clinicians viewed these data in 81.9% (158/193) of these visits. Overall user-friendliness on the System Usability Scale received a median rating of “good” from both patients and clinicians. Resources included costs to build the data transfer pipeline, a 24-month build timeline, and ongoing technical support.

**Conclusions:** By combining user-centered design principles with dedicated resources, we built a feasible and usable digital health solution to bridge the gap between brief clinic visits and daily life in outpatient rehabilitation. These results and future refinements will facilitate broader-scale implementation of sensor technology in rehabilitation.

## INTRODUCTION

The primary goals of patients seeking outpatient neurorehabilitation care are to improve their activity in daily life.^1, 2^ Rehabilitation clinicians’ ability to help patients achieve these goals is limited by the episodic nature of clinic visits, which provide only brief, intermittent contact with patients. This model of care often requires clinicians to infer patients’ activity in daily life from limited or indirect information. In standard rehabilitation practice, the two primary approaches clinicians use to estimate patients’ activity in daily life are in-clinic assessments and patient self-report measures. When in-clinic assessments are used, the assumption is that scores on these assessments translate to activity in daily life. For example, a clinician might use scores on an in-clinic test of walking endurance (e.g., the 6-Minute Walk Test^3^) to estimate their patient’s walking activity in daily life. This approach can lead the clinician to focus their treatment plan and goals on improving the patient’s score on the in-clinic walking endurance test,^4^ under the assumption that improvements on this test will translate to more steps taken in daily life. However, recent research indicates that this assumption frequently does not hold.^5–7^ When self-report measures are used, the assumption is that patients can reliably recall and accurately estimate their activity. Studies have shown, however, that recall bias and over- and under-reporting can greatly limit the utility of self-report measures in estimating a person’s actual activity in daily life.^8–10^ Indeed, self-reported measures of activity can vastly differ from direct (sensor-based) measures.^8–11^ Collectively, these data indicate that clinicians cannot rely solely on in-clinic assessments or a patient’s self-report of their activity to accurately estimate their patients’ activity in daily life. If clinicians do not have access to reliable data on how much their patients are moving in daily life, they cannot effectively intervene to change it.

One technology with strong potential to bridge the disconnect between the clinic and daily life is wearable sensing. In the research realm, wearable sensors are becoming commonplace, offering a reliable approach to both measure and improve daily life outcomes. Studies have shown that accelerations from wrist-worn sensors can be translated into variables that reflect upper limb use in daily life.^12–15^ Other studies demonstrate the reliability and validity of wrist, hip, or ankle-worn sensors for measuring step counts and identified key movement characteristics that affect step count algorithms in rehab populations.^16–19^ Studies also indicate that when patients and clinicians have access to wearable sensor data, patient outcomes can improve. For example, a recent randomized controlled trial found that providing activity data from a wearable sensor to people with stroke and their physical therapist resulted in improvements in activity in daily life (steps/day).^6^ This study, among others,^20, 21^ highlights the potential of wearable sensor technology to bridge the gap between brief, in-clinic visits and daily life by providing reliable and actionable data from daily life.

Despite this potential, wearable sensors are not part of routine rehabilitation care.^22–25^ Several factors may explain the lack of routine use in rehabilitation practice, including limited awareness of the disconnect between in-clinic assessments and activity in daily life and/or limited understanding of the capabilities of wearable sensor technology. To explore these factors, our preliminary work employed a mixed-methods design to examine the perspectives of patients and clinicians.^26^ We found that clinicians (physical and occupational therapists) and patients acknowledged the disconnect between the clinic and daily life and recognized wearable sensor technology as a potential solution to address this disconnect. Clinicians and patients also expressed preferences for how these data could best be integrated into routine clinical care.^26^ Collectively, these findings highlighted a clinic-to-daily-life practice gap acknowledged by patients and clinicians, a potential solution to address this gap, and stakeholder preferences for integrating the solution into clinical care.

Guided by these findings, the purpose of the current study was to develop and test a digital health solution that integrated wearable sensor technology into routine rehabilitation practice. The goal was to develop a solution that would work within existing clinical workflows while minimizing burden on patients and clinicians. To achieve this goal, we used a user-centered design approach that included iterative feedback from patients and clinicians throughout the development and testing of the digital health solution. The feasibility, usability, and resources required of the solution were then evaluated in a proof-of-concept study in outpatient rehabilitation. Feasibility was quantified using the following outcomes: clinician and patient engagement in the program, patient adherence to daily data syncing, sensor data availability at the time of clinic visits, and whether clinicians viewed the sensor data. We also evaluated the relationship between patients’ self-reported digital technology self-efficacy and adherence to daily syncing and hypothesized that higher digital technology self-efficacy would be associated with higher syncing adherence. Usability was measured quantitatively by the System Usability Scale, in which we hypothesized that clinicians and patients would report that the system is usable, and qualitatively by using clinician- and patient-specific surveys to collect general feedback on their experiences using the program. Resources quantified included the costs, time, personnel, and tech support needed to build and maintain the program.

## METHODS

This study used a user-centered design approach to develop and refine a digital health solution to address the disconnect between clinic visits and daily life, which was then evaluated in a proof-of-concept study in outpatient rehabilitation care. The Washington University Human Research Protection Office approved the study. All participants provided written informed consent to participate in the study.^27, 28^

### User-centered Design of a Wearable-to-EHR Digital Health Solution

User-centered design is the process of engaging end users early and often in the design and development of digital health solutions.^29–31^ This is key as digital health initiatives are more likely to be successful if stakeholders are engaged early and often.^31–33^ Figure 1 displays the user-centered design process that guided the development of the digital health solution.

**Figure 1.**
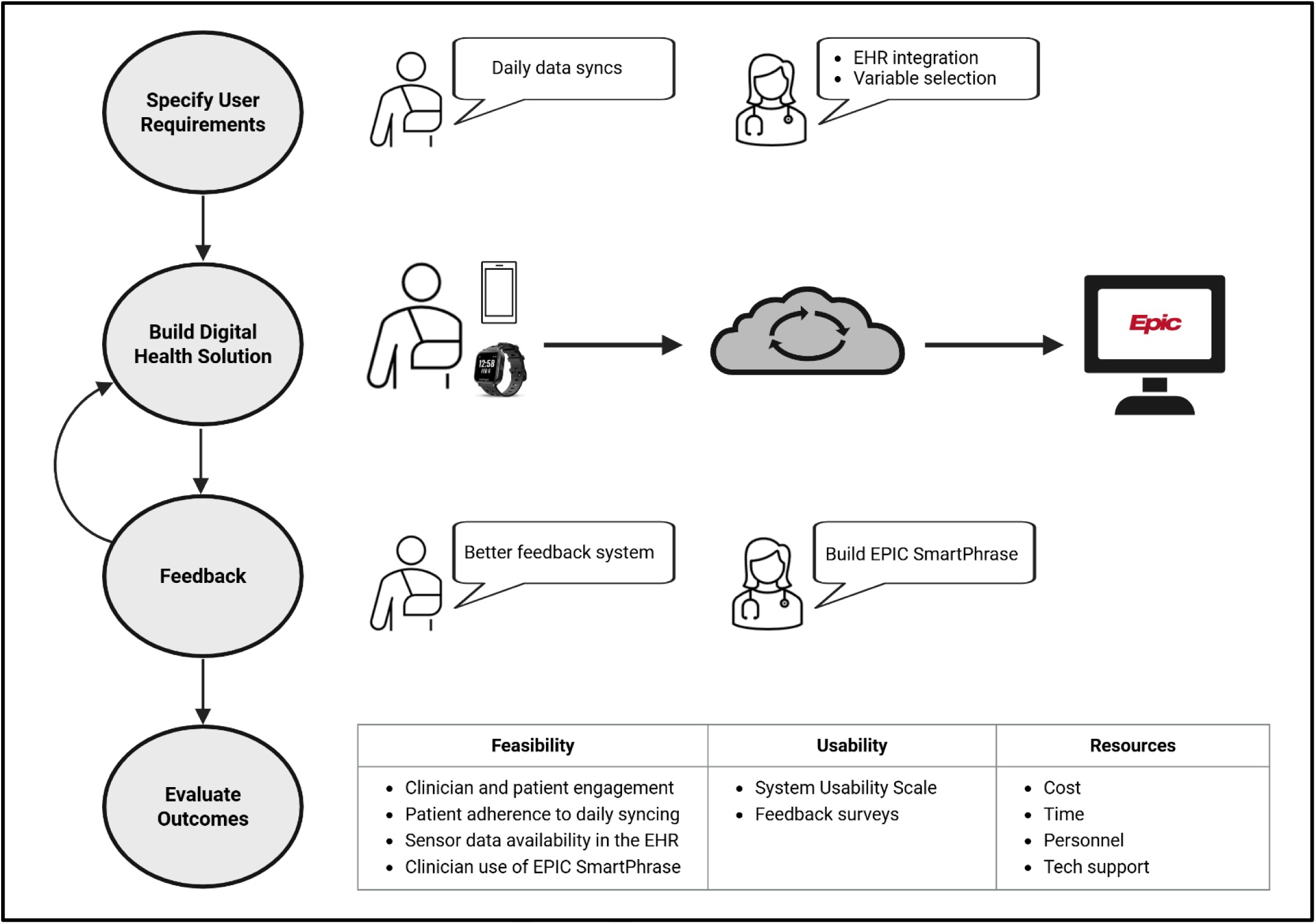
Development and Evaluation of the Wearable-to-EHR Solution. The left side of the figure shows the user-centered design steps undertaken to develop and evaluate the wearable-to-EHR solution. “Specify User Requirements”: This step was accomplished in our preliminary mixed methods work, in which patients expressed a preference for no greater than daily data syncs, and clinicians expressed preferences for integration of specific sensor variables into the EHR. “Build Digital Health Solution”: A data transfer pipeline was built using the Ametris LEAP sensor worn on the patient’s wrist. The patient’s treating clinician or a member of the research team assisted the patient with downloading the CentrePoint Connect app. Patients were instructed to press a “manual sync” button in the app once per day, which allowed raw data to be transferred from the sensor to the CentrePoint cloud for storage and processing into the summary variables specified by the clinicians in the prior step. Summary variables were transferred from the CentrePoint cloud into a flowsheet in Epic via the CentrePoint application programming interface (API) each weekday morning. “Feedback”: After initial testing, patients provided feedback to improve the frequency and approach in which they would receive their sensor data. Clinicians provided feedback to build an Epic SmartPhrase to facilitate data viewing in the EHR. Where possible, this feedback was incorporated into the digital health solution build. “Evaluate Outcomes”: This table displays the feasibility, usability, and resource outcomes evaluated in the proof-of-concept study.

### Specify User Requirements

The goal of this first step, which was accomplished in our preliminary work, was to understand patient and clinician awareness of the clinic-to-daily life gap and preferences for how wearable sensor technology could be integrated into clinical care as an opportunity to address this gap.^26^ Data from this mixed-methods study revealed acknowledgement of the gap and wearable sensor technology as a potential solution to address it. Clinicians (physical and occupational therapists) and patients also had preferences for how the technology could most seamlessly integrate into clinical workflows while minimizing burden. A key preference voiced by clinicians was integration of sensor data into the electronic health record (EHR) to minimize the number of external applications that would need to be accessed to view the patient’s sensor data. This was anticipated as integrating remotely collected data within the EHR is a commonly voiced preference among providers in other areas of digital health research.^32, 34–36^ Clinicians also had preferences for specific sensor variables to be available in the EHR. These included step counts, sleep hours, heart rate, and minutes spent at various intensity levels (e.g., sedentary, light, and moderate-to-vigorous). For sensor-to-EHR data transmission, patients preferred a low-burden, user-friendly data syncing process at a maximum frequency of once per day.^26^ These preferences were leveraged in the second step to build a wearable-to-EHR digital health solution.

### Build Digital Health Solution

The solution was built around the Ametris LEAP wrist sensor (Pensacola, FL) and Epic (Epic Systems Corporation),^37^ WashU’s EHR system. The Ametris LEAP sensor was selected for its reliability and validity in measuring activity in daily life,^38–41^ its heart rate monitoring capabilities desired by clinicians,^26^ and its customizable companion cloud platform, CentrePoint, which permitted remote and automated data uploads via the CentrePoint Connect mobile app. Raw sensor data were automatically transmitted via Bluetooth to the mobile app whenever the sensor was in range of their phone, aligning with patient preferences for low-burden syncing. Raw data were then transmitted from the mobile app to the CentrePoint Cloud via a cellular or Wi-Fi signal where they were stored and processed into summary variables before entering the EHR. The summary variables were selected based on clinician preferences gleaned in the prior step and included sleep hours, step counts, heart rate (median), heart rate (5^th^ and 95^th^ percentiles), sedentary minutes, light intensity minutes, moderate-to-vigorous intensity minutes, and hours of upper limb use (for the upper limb on which the sensor was worn).^39, 42^ Ametris custom-engineered their CentrePoint platform to integrate the hours of upper limb use algorithm. This algorithm used accelerations from the upper limb on the side the sensor was worn to compute the number of hours of upper limb movement and was made available as a potentially useful variable for patients with goals to improve their upper limb activity.^13, 43^ The summary variables were pulled from the CentrePoint cloud into a flowsheet in Epic each workday morning via Ametris’ CentrePoint API (application programming interface). The Epic flowsheet was organized such that each column was a calendar date and each row was a sensor variable (Supplemental Figure 1). Clinicians could view their patient’s sensor data in Epic by navigating to the flowsheet tab. Patients could optionally view their sensor data in the clinician’s note in MyChart (Epic Systems Corporation),^44^ Epic’s patient-facing portal, but could only do so after a clinic visit and if sensor data were documented in the clinician’s note. The solution was designed to work best with daily syncing to ensure continuous data flow from sensor to Epic. If a sync was missed on a particular day, however, data were not lost but their availability in Epic was delayed and subsequent syncing times increased.

### Feedback

The wearable-to-EHR solution was initially piloted by one physical therapist and two patients to collect feedback and identify areas for refinement. The physical therapist recommended building an Epic SmartPhrase that would allow clinicians to pull sensor data from the flowsheet into their clinical note to maximize viewability and usability of the data. This would eliminate the need for clinicians to leave the clinical note and access the flowsheet tab in Epic to view sensor data. Because this feedback was straightforward to implement, it was incorporated quickly. A SmartPhrase was built that pulled the patient’s most recent 16 days of sensor data into the clinician’s note. The primary feedback voiced by patients during pilot testing was to improve the timeliness and ease of access to sensor data. The patients reported that accessing MyChart could be cumbersome and may be especially burdensome for those who do not regularly access MyChart after a health care encounter. This feedback was shared with WashU informaticians. Although plans for a new patient feedback pipeline were developed, implementation and testing would require additional time. Therefore, this modification was not introduced immediately, and the study continued while the new pipeline was being developed. Accordingly, data reported here reflect the patient feedback pipeline delivered through MyChart. Pilot testing also revealed that automated syncing frequently led to incomplete data transfer from sensor to Epic. Subsequent testing and consultation with Ametris and WashU informaticians confirmed that manual in-app syncing resolved the issue. Thus, future patients were instructed to avoid relying on automated syncing and instead press a “manual sync” button in the CentrePoint Connect app once per day to ensure complete data transfer. Thus, the study proceeded with a new Epic SmartPhrase, building of a new patient feedback pipeline (not incorporated immediately), and transition from automated to manual syncing.

### Evaluate Outcomes

Three categories of outcomes were then evaluated in a proof-of-concept study: *feasibility*, *usability*, and *resources*. *Feasibility* outcomes included the following:

• Clinician and patient engagement in the program: This was quantified as the number of clinicians and patients recruited over the 9-month study timeframe, the number of clinicians who trialed the program with more than one patient, and the average duration of monitoring.

• Patient adherence to daily syncing: Adherence to daily data syncing was calculated for each patient as the number days the patient successfully synced their data relative to the total number of days monitoring, expressed as a percentage. If a patient performed only an incomplete sync on a particular day, this day was not counted as a successful syncing day. If an incomplete sync was preceded or followed by a complete sync within the same day, however, this day was counted as a successful syncing day. Syncing adherence was calculated at the group level using descriptive statistics. Pearson’s correlation was used to evaluate the relationship between syncing adherence and digital technology self-efficacy, measured by the Digital Technology Self-Efficacy Survey,^45–47^ which was completed by participants at the start of the study. Participants rated how much they agreed or disagreed with 17 statements on a 4-point Likert scale, ranging from “strongly disagree” to “strongly agree”. Examples of statements include, “I find working with digital technology very easy” and “I find it difficult to get digital technology to do what I want it to”. Responses to each item were translated to a numeric value between 1 (low digital technology self-efficacy) and 4 (high digital technology self-efficacy) and then summed to provide a total score that ranged between 17 and 68. A statistically significant (*p* < 0.05) Pearson’s correlation indicated our hypothesis was met.

• Sensor data availability in Epic: Recent sensor data were classified as “available” if they were present in Epic within 3 days prior to the clinic visit and “unavailable” if the most recent data in Epic were more than 3 days old. A 3-day window was chosen to account for expected data transfer delays over the weekend when data were not processed in the CentrePoint cloud or pulled into Epic. When calculating clinic visits, we excluded the visit where the sensor was first issued and the very last visit where use of the sensor was discontinued.

• Clinician viewing activity in Epic: Clinician review of sensor data was determined by use of the associated SmartPhrase in the clinical note. If the SmartPhrase was used in the clinical note, data were considered to be viewed. We then calculated the number of clinic visits within the monitoring period where data were viewed by the clinician in relation to the recency of sensor data in Epic.

#### Usability

Usability was measured quantitatively using the System Usability Scale (SUS) which was completed by clinicians and patients at the end of their participation in the study. The SUS is a validated and commonly used 11-item survey administered to end-users of digital health initiatives.^48–51^ The first 10 items asked participants to report how much they agreed with a particular statement (e.g., “I thought the system was easy to use”) using a 5-point Likert scale ranging from “strongly disagree” to “strongly agree”. The 11^th^ item asked participants to provide an overall ranking of the system on a 7-point Likert scale ranging from “worst imaginable” [1] to “best imaginable” [7].^51^ We computed the frequency of responses for items 1-10 of the SUS to identify strengths and areas of improvement for future work. To test our hypothesis that clinicians and patients would report that the system was usable, we analyzed responses to the 11^th^ item regarding their overall ranking of the system.^51^ We operationalized a useable system as one in which the median score of both clinician and patient cohorts was at least “good” [5].^52^ Usability of the wearable-to-EHR solution was evaluated qualitatively using patient and clinician feedback surveys. The purpose of these surveys was to gain information on the strengths and areas for improvement that were not captured by the SUS. The survey items were initially developed by the research team and refined based on input from patients and clinicians to ensure key elements were captured. Patients and clinicians completed these surveys approximately 3-4 weeks after onset of using the technology.

#### Resources

We quantified the following costs, time, personnel, and tech support needed to build and maintain the wearable-to-EHR solution:

- Costs: Data transfer pipeline build, consultation fees for regulatory support, and purchase of sensors and wear accessories (wrist straps and chargers).
- Time: Time to build data transfer pipeline, number of group in-services and individual training sessions requested by clinicians to assist their patients with setting up the sensor.
- Personnel: Number of internal and external partners that were involved to approve, build, and manage the data transfer pipeline.
- Tech support: Number of tech support cases handled by research staff, number of text message reminders sent by research staff to patients who had not synced their sensor data in ≥ 3 days.

Resources in the cost, time, and personnel domain were tracked in an Excel document located on WashU’s secure Box server. Tech support requests and syncing reminders were tracked in REDCap (Research Electronic Data Capture).^53, 54^ Statistical analyses were conducted in R (R Core Team 2022, version 4.2.1) and included use of the tidyverse package.^55, 56^

### Proof-of-Concept Study Procedures

Clinicians and patients were recruited from two outpatient rehabilitation clinics within the WashU Medicine system between November 2024 and July 2025. Our goal was to recruit at least 10 clinicians and 15 patients as these are typical recruitment targets of early phase studies in the digital health literature.^57–60^ Clinicians were eligible to participate if they were a licensed physical or occupational therapist or assistant and employed at WashU. Clinicians were primarily recruited via in-services delivered by the research team and targeted emails. Patients were eligible to participate if they were at least 18 years of age, being seen for outpatient rehabilitation services at WashU, and had access to a mobile phone or tablet for data syncing. Patients were recruited through clinician referral. Clinicians were encouraged to refer any eligible patients for whom they felt having access to their sensor data could be informative for the patient’s care. Patient eligibility criteria were intentionally broad to mirror the wide heterogeneity seen in routine outpatient rehabilitation care. Upon enrollment, demographic information was collected from clinicians via survey and from patients via the medical record (Epic). Clinicians and patients also rated the importance of measuring activity outside the clinic on a 4-point Likert scale ranging from “not important” to “very important”.

Each clinic received an inventory of sensors, wrist bands of various sizes, and chargers. The research team trained participating clinicians on how to issue the sensor to their patients and view sensor data in Epic. Training consisted of group in-services and individual training sessions. Clinicians were provided with a manual containing detailed instructions that they could refer to as needed throughout the study. As clinicians referred patients to the study, they indicated whether they would like a research team member to be present at the patient’s clinic visit to assist with setting up the sensor. During the visit, the clinician and/or research team member provided the patient with a sensor and charger and assisted them with installing the CentrePoint Connect app on their mobile phone or tablet to permit data syncing. Patients were instructed to sync their data in the mobile app once per day^26^ and charge the sensor every 3-4 days to avoid reaching its 7-day battery life. If a day of syncing was missed, patients were instructed to sync as soon as possible to facilitate data transfer into Epic and minimize future syncing times. Patients were given a handout that described the syncing, charging, and data viewing procedures as well as the contact information of the research team for any questions about the study and/or technical assistance with using the technology. The duration of monitoring was left to the discretion of the clinician and patient. Monitoring could continue for any length of time prior to the patient discharging from outpatient services so long as the clinician and/or patient found the information valuable and the patient was willing to wear the sensor and sync their data. The sensor was collected prior to patient discharge, cleaned, and placed back in a sensory inventory station to be used with other patients.

## RESULTS

### Feasibility

This analysis includes all clinicians and patients who consented into the study and who completed monitoring over the 9-month study period. During this time, 17 clinicians consented into the study and referred a total of 43 patients. Of the 43 referred patients, 4 declined to participate or did not respond to the research team’s request to complete the informed consent, yielding 39 patients who consented to participate. One of the 39 patients did not return to outpatient rehabilitation after the initial visit and was excluded from the analysis. This resulted in 38 patients who completed monitoring within the 9-month period (average of 4 patients/month consented) and were included in the analysis. Tables 1 and 2 display the demographic characteristics of the clinician and patient cohorts, respectively.

**Table 1.** Clinician Demographics (n =. 17)

|  |  |  |
| --- | --- | --- |
| **Age (years)** |  | 30 [27.0, 36.0] |
| **Sex** | Female<br>Male | 94.1% (16)<br>5.9% (1) |
| **Race** | White<br>Asian | 94.1% (16)<br>5.9% (1) |
| **Profession** | Physical therapist<br>Physical therapist assistant<br>Occupational therapist | 76.5% (13)<br>5.9% (1)<br>17.6% (3) |
| **Education** | Masters/Doctorate degree<br>Bachelors degree | 88.2% (15)<br>11.8% (2) |
| **Total number of years practicing** |  | 6 [2.5, 10.0] |
| **Number of years practicing at current setting** |  | 3 [2.0, 5.5] |
| **Years of experience using wearable motion sensors in clinical practice** | None<br><1 year<br>1-2 years<br>3-5 years<br>≥6 years | 11.8% (2)<br>11.8% (2)<br>35.25% (6)<br>35.25% (6)<br>5.9% (1) |
| **How important is your ability to monitor your patient's activity outside the clinic?** | Very important<br>Somewhat important<br>Neutral<br>Not important | 76.5% (13)<br>23.5% (4)<br>0% (0)<br>0% (0) |

Continuous variables are reported as median [1<sup>st</sup> quartile, 3<sup>rd</sup> quartile]. Categorical variables are reported as percentage (count).

**Table 2.**
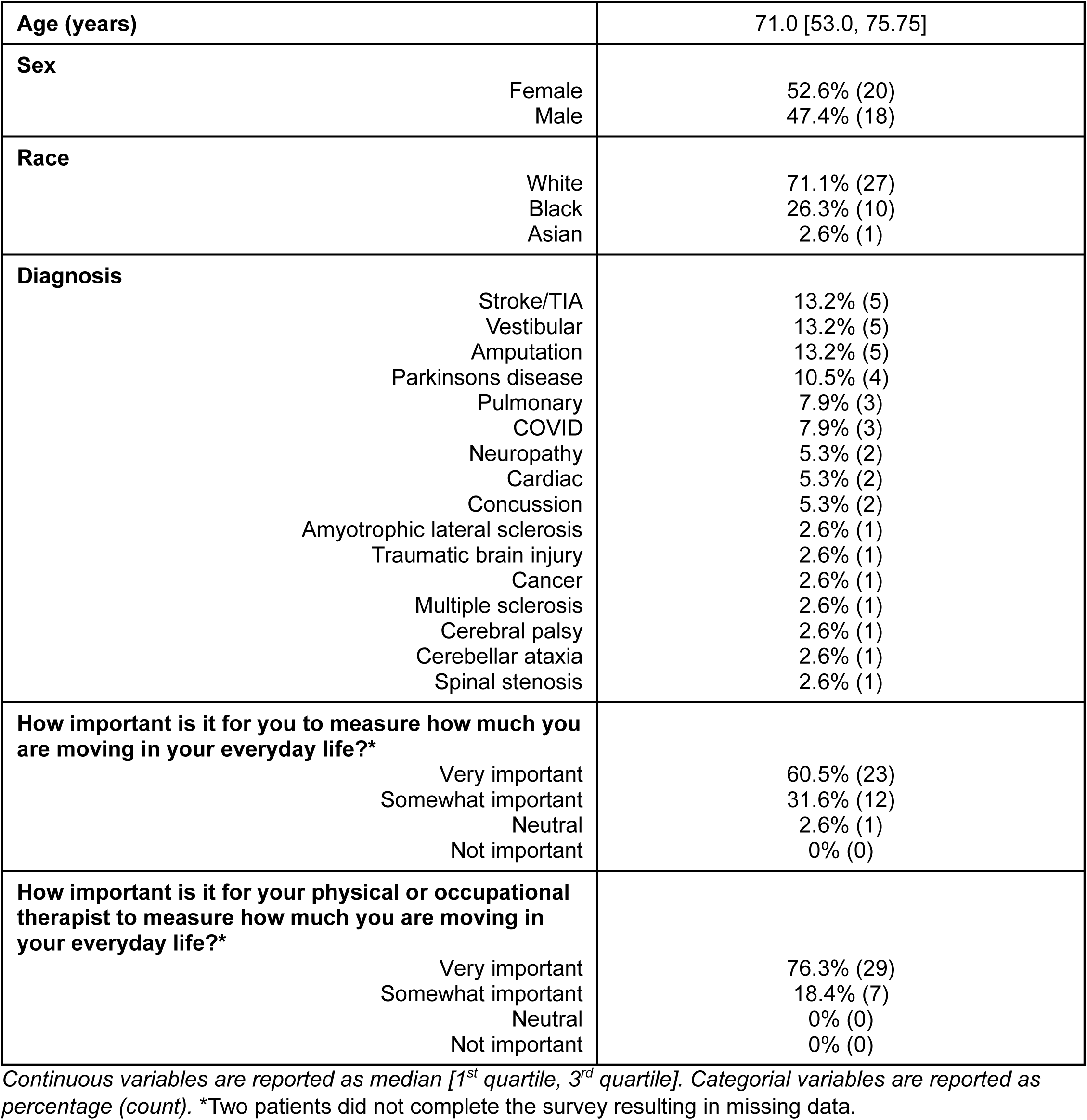
Patient Demographics (n = 38)

Ten clinicians trialed the program with more than one patient (Figure 2A). For the patient cohort, the median [1^st^ quartile, 3^rd^ quartile] length of monitoring was 42.0 days [30.25, 72.0] (Figure 2B). Adherence to daily syncing was 68.6% [50.0%, 87.05%]. Digital technology self-efficacy was generally higher in the clinician cohort compared to the patient cohort but was highly variable within cohorts (Figure 2C). This was to be expected as some individuals reported being more “tech savvy” than others based on their past experiences and occupational requirements. For example, one patient reported working in their employer’s IT department and likely had more opportunities to interact with technology than others. Conversely, several patients preferred a “hands-off” approach to technology and used their partner’s phone/tablet for data syncing due to lack of confidence or physical or cognitive limitations. Contrary to our hypothesis, digital technology self-efficacy was unrelated to syncing adherence in the patient cohort (Pearson’s *r* = 0.03, *p* = 0.86; Figure 2D).

**Figure 2.**
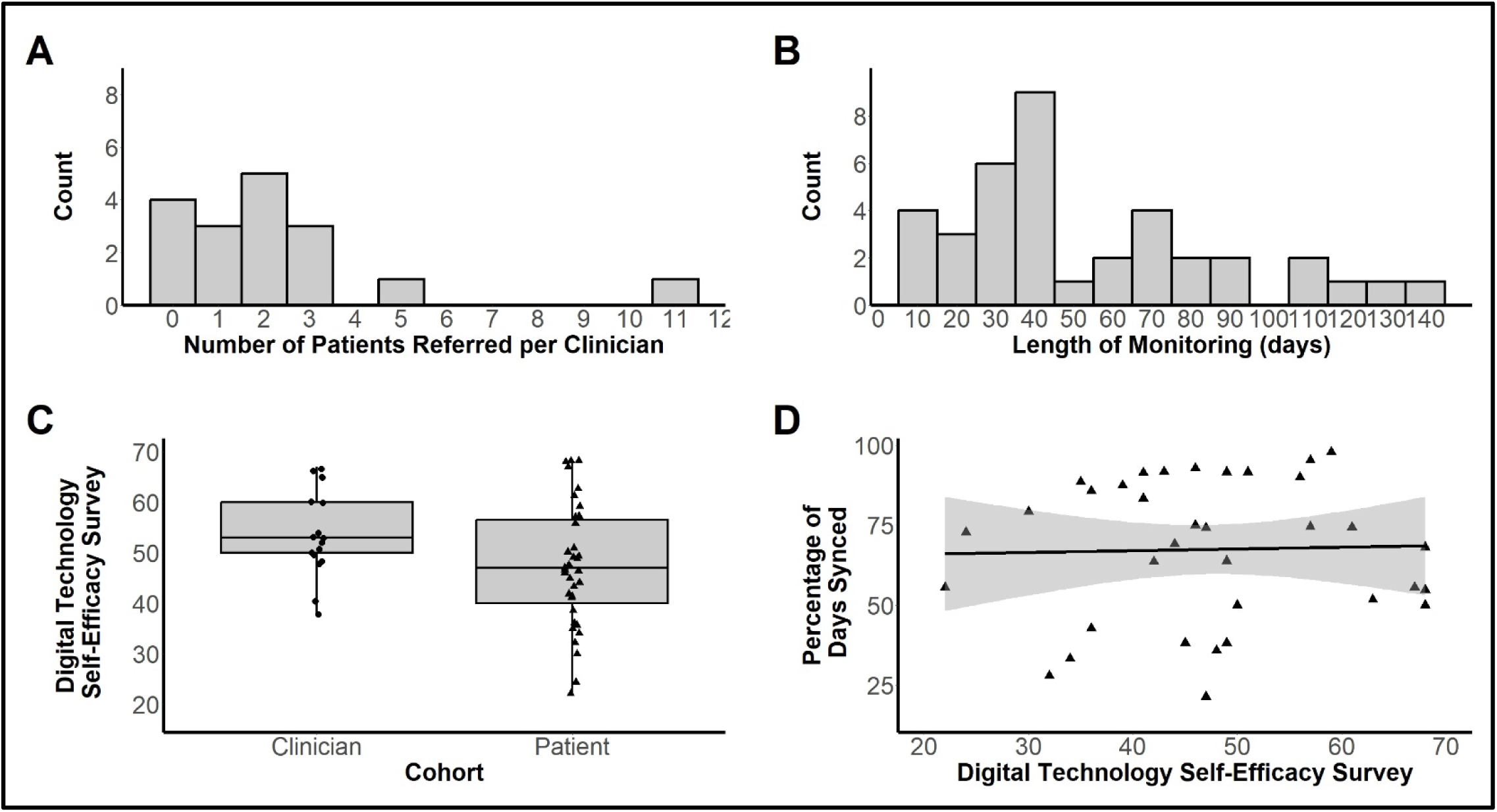
Feasibility Outcomes. **A-** Histogram showing the number of patients (X-axis) referred by clinicians (Y-axis), e.g., 3 clinicians referred 1 patient each. **B-** Histogram of the number of days of monitoring across patients. **C-** Box plot showing results of the Digital Technology Self-Efficacy Survey for the clinician and patient cohorts. **D-** Scatter plot showing the lack of relationship between the percentage of days synced and the Digital Technology Self-Efficacy Survey for the patient cohort. The black line shows Pearson’s correlation (r = 0.03, p = 0.86), and the shaded region shows the 95% confidence interval.

Across the 38 patients, 214 clinic visits occurred. Recent (≤ 3 days) sensor data were available in Epic for 193 (90.2%) of these clinic visits. Of these 193 clinic visits, the SmartPhrase was used in the clinical note on 158 (81.9%) visits, indicating data were viewed. The SmartPhrase was not used in the clinical note for the remaining 35 (18.1%) visits. Of the 21 (9.8%) clinic visits in which the patient did not perform a recent sync and recent data were therefore not available in Epic, the SmartPhrase was not used in 8 notes but was used in 13 notes. The presence of the SmartPhrase in these 13 notes in the absence of recent data suggests the clinician was interested in viewing the data despite recent data not being available at the time of the visit.

### Usability

Overall, patient and clinician responses to items 1-10 of the SUS were highly variable (top part of Figure 3). Items pertaining to the complexity of the system as well as the learning required to get going with the system tended to have more favorable ratings by patients and clinicians. Items pertaining to the integration of the various functions in the system and overall inconsistency in the system had more variable responses, suggesting these may be areas for improvement. The median score for the overall user-friendliness of the system was “good” [5] for both the patient and clinician cohorts, indicating our hypothesis was met (bottom part of Figure 3).

**Figure 3.**
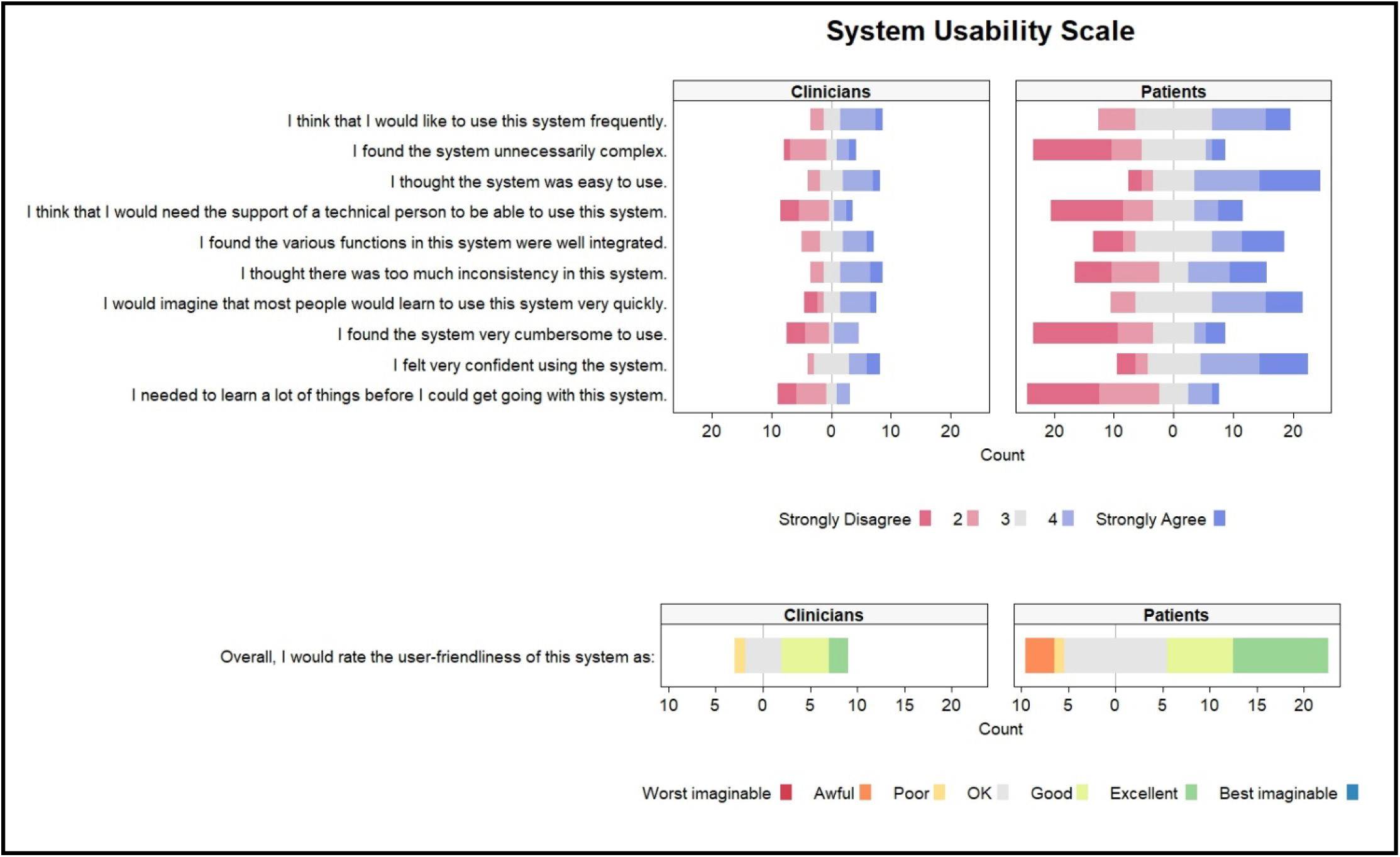
System Usability Scale Results. The top portion of the figure shows the responses to items 1-10, which are scored on a 5-point Likert scale ranging from “strongly disagree” (dark red) to “strongly agree” (dark blue). The bottom portion shows the responses to item 11, which is scored on a 7-point Likert scale ranging from “worst imaginable” to “best imaginable”. The width of each colored section of each bar represents the number of individuals who endorsed the response (e.g., 12 patients endorsed the response “strongly disagree” to the question, “I needed to learn a lot of things before I could get going with this system.”). Clinician and patient responses are shown in the left and right panels, respectively.

Additional insights were gleaned from the clinician and patient feedback surveys. In general, responses to most items on the clinician survey were positive (top part of Figure 4). Most or all clinicians reported processes related to issuing the sensor, locating sensor data in Epic, and using the SmartPhrase were easy. Critically, most clinicians reported that the information from the sensor added value to their patient care. Clinicians expressed mixed views on the time spent providing tech support to patients, highlighting a potential area for improvement. In general, most patients reported that the sensor was comfortable to wear, charging the sensor was easy, and sensor data were being discussed with their clinician (bottom part of Figure 4). Items that had more variable responses for patients were ease of syncing and perceived usefulness of sensor data for themselves. Querying patients further on this item revealed that patients did not prefer viewing their data in the clinician note in MyChart, providing statements such as “I forgot it was in there” and “I can’t find it in MyChart”.

**Figure 4.**
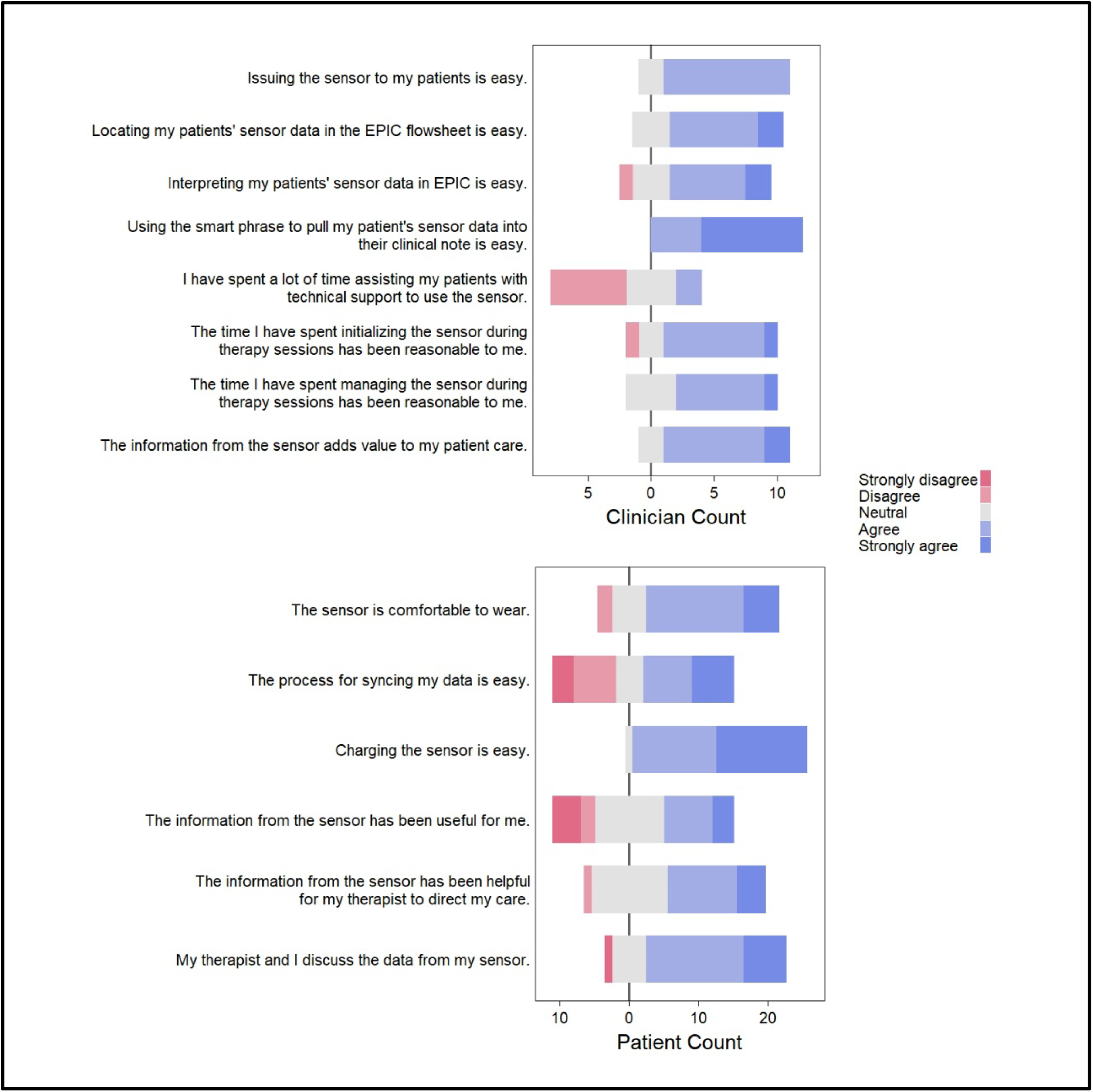
Clinician and Patient Feedback on the Wearable-to-EHR Solution. Participants responded to all items on a 5-point Likert scale ranging from “strongly disagree” (dark red) to “strongly agree” (dark blue). The width of each colored section of each bar represents the number of individuals who endorsed the response (e.g., 6 clinicians endorsed the response “disagree” to the question, “I have spent a lot of time assisting my patients with technical support to use the sensor.”). Clinician and patient responses are shown in the top and bottom panels, respectively.

### Resources

Figure 5 provides a schematic of the resources (costs, time, personnel, and tech support) that were required to build and maintain the program. Costs fell into four categories: data infrastructure build, data storage and license fees, consultant fees, and purchase of sensors, chargers, and wear accessories (i.e., wristbands). The total cost to build the data transfer infrastructure ($24,541) included custom work from Ametris to integrate the hours of upper limb use algorithm into their CentrePoint Cloud platform and modify their API to support data integration into Epic. In the time domain, the infrastructure build took 24 months, with an additional 4 months required to train all clinicians as they identified and referred patients on their caseloads. On average, clinicians requested two individual sessions with a research staff member present at the patient’s clinic visit to assist with issuing the sensor prior to feeling independent with the procedures and requiring only remote tech support as needed. In the personnel domain, six internal groups were involved, reflecting the various legal, privacy, and security approvals required before building the wearable-to-EHR solution. In the tech support domain, 28 out of 38 patients (73.7%) required at least one text message reminder to sync their data. The median number of reminders for these 28 patients was 2 [1.0, 3.5], with one patient requiring 9 reminders. The research team troubleshooted 46 tech support requests from 15 clinicians and 8 patients. Technical issues were first addressed over the phone and escalated to a home visit if necessary.

**Figure 5.**
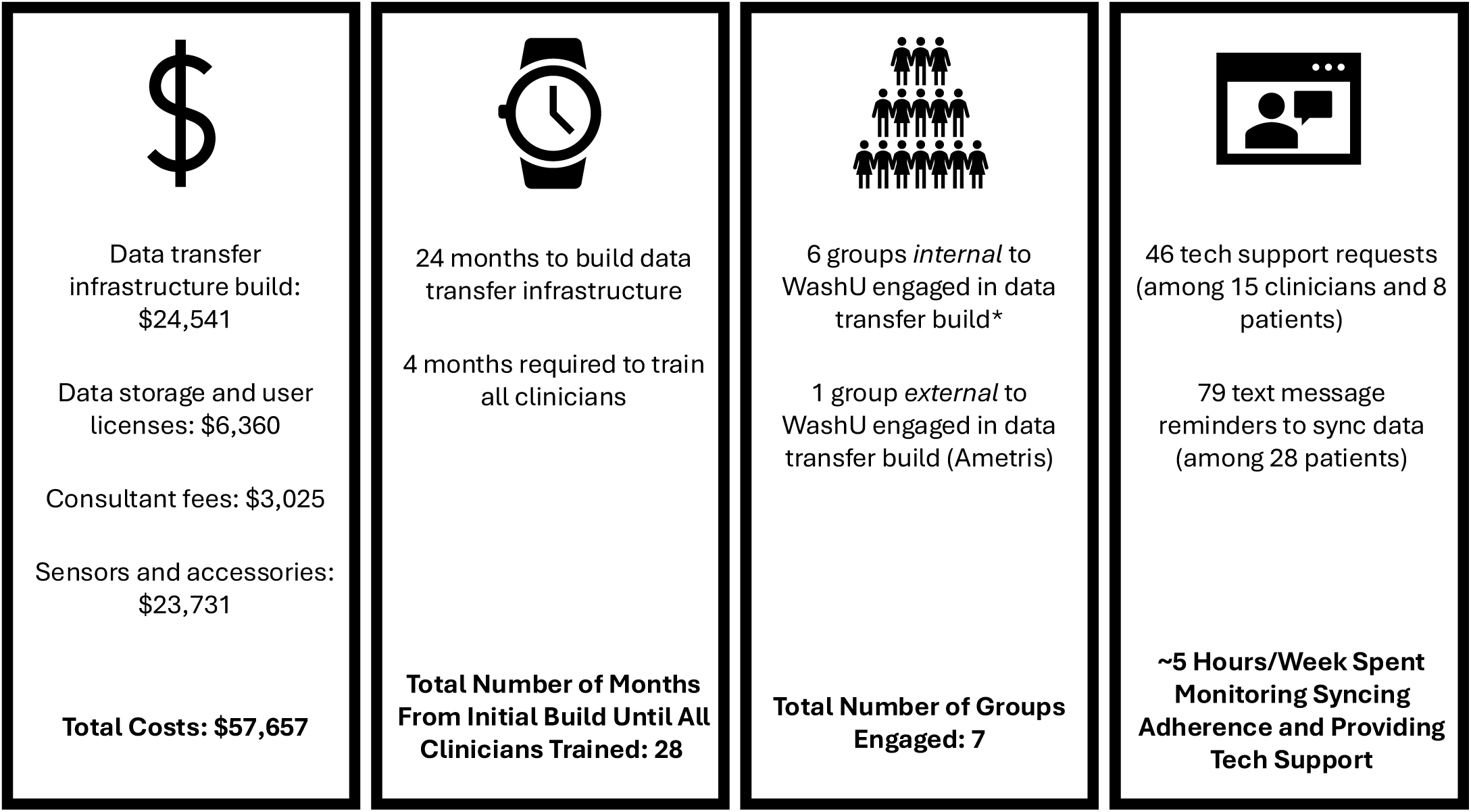
Resources Required to Build and Maintain the Wearable-to-EHR Program. From left to right, the panels display the costs, time, personnel, and tech support required to build and maintain the wearable-to-EHR solution. *Internal groups engaged: HIPAA Privacy Office, Joint Research Office for Contracts, mHealth Research Core, Office of Information Security, Office of the Vice Chancellor for Research, and the Institute for Informatics, Data Science, and Biostatistics.

## DISCUSSION

This study used a user-centered design approach to develop and refine a wearable-to-EHR digital health solution, and then evaluated its feasibility, usability, and resource requirements in a proof-of-concept study in outpatient rehabilitation care. A user-centered design approach allowed us to build a solution collaboratively with patients and clinicians to optimize its integration within clinical workflows and minimize end-user burden. Data from the proof-of-concept study indicate that the wearable-to-EHR solution was feasible and usable in clinical practice and required resources to build and maintain. Collectively, these data support the potential of wearable sensor technology to bridge the gap between the clinic and daily life, highlight the importance of critically evaluating feasibility, usability, and resource outcomes in the development of digital health solutions, and lay the groundwork for future studies to investigate the impact of these data on clinical practice and patient outcomes.

A key, but often overlooked, catalyst that contributed to the study’s positive findings was building a digital health solution that addressed a problem of direct importance to both patients and clinicians. This was accomplished in the first step of our user-centered design approach that confirmed the clinic-to-daily-life gap was salient to clinicians and patients. If end users of a digital health solution do not value the problem it seeks to address, the solution is more likely to fail.^32, 61^ The user-centered design approach also allowed us to identify key data collection preferences of patients’ and clinicians’ that were critical for the wearable-to-EHR build and likely facilitated high engagement in the program. The implications of not understanding these preferences include development of a solution ill fit for clinical workflows, greater end-user burden, and resources wasted to build a solution misaligned with end-user preferences and therefore unlikely to be sustainable. As digital health becomes integral to modern rehabilitation practice, this study and others^62–64^ highlight the essential role of engaging end-users early and often throughout the development and testing of digital health solutions.

Data from this proof-of-concept study provide evidence for feasibility of the wearable-to-EHR solution in outpatient rehabilitation clinical practice. Steady and repeat referrals from clinicians yielded four consented patients per month, allowing us to exceed our recruitment targets. This, along with the 42-day median duration of monitoring, suggests that, despite prior reports of low utilization,^22–25^ clinicians and patients are willing to use, or at least trial, new technologies in rehabilitation clinical care. Although 4 of the 17 consented clinicians did not refer any patients during the 9-month study period, each has since referred patients as recruitment has continued. This willingness to engage with the program was also seen in patient syncing adherence and clinician viewing data. A median syncing adherence of 69% demonstrates strong preliminary feasibility, given the proof-of-concept nature of this study and the fact that daily syncing became a manual process. It is also important to note that although daily syncing was encouraged to facilitate continuous data transfer, it wasn’t always necessary. For example, if a patient missed a data sync on a Monday but successfully synced on Tuesday, recent data in Epic should still be available for a Wednesday clinic visit. Despite this, daily syncing adherence varied widely, with some patients syncing on only ∼25% of days. Contrary to our hypothesis, individuals who were less adherent to daily syncing didn’t necessarily have lower digital technology self-efficacy (Figure 2D). The low correlation indicates that individuals with lower digital technology self-efficacy were still able to successfully participate in the program and that higher digital technology self-efficacy is neither a requirement for nor a guarantee of consistent daily syncing. The variability in daily syncing adherence, coupled with the frequent need for reminders and technical support (discussed below), underscores the importance of low-burden, user-friendly syncing procedures and dedicated tech support resources.

Feasibility of the wearable-to-EHR solution was also demonstrated by the fact that recent data were available in Epic for 90.2% of clinic visits and viewed (through use of the SmartPhrase) in 81.9% of those visits. Thus, for most clinic visits, recent data were available and viewed by the clinician. This justifies the investment of resources to deliver sensor data directly into the clinicians’ preferred location (Epic). The absence of a recent data sync prohibited data from being available in Epic for the remaining 9.8% of visits. These findings reinforce the importance of assessing the critical actions of patients (data syncing) and clinicians (data viewing) separately as they can lead to different interventions. Low syncing adherence requires intervention at the patient-level, such as reminder notifications, adding automations where possible, and ensuring rapid-response and user-friendly tech supports are available. Conversely, low viewing activity would require intervention at the clinician-level, such as notifications in the EHR to alert clinicians to the presence of new data or placing data in a more seeable location. If this work had initially focused on the utilization of sensor data in clinical practice as the outcome, these critical insights would have been missed. These feasibility data provide evidence that patients and clinicians can perform their critical actions (data syncing by patients, data viewing by clinicians) to make the wearable-to-EHR solution viable.

Data from the SUS and feedback surveys demonstrated that the wearable-to-EHR system was usable and identified strengths and areas for improvement. Achieving an overall rating of “Good” on the SUS is common target in the digital health literature and allowed us to benchmark the usability of the wearable-to-EHR solution against others.^52^ Despite achieving this target, the variability in responses to items pertaining to the integration and consistency of various functions in the system indicate areas for improvement in the future. We instructed patients and clinicians to consider all tasks related to their interaction with the solution when completing the SUS. Consequentially, this limited our ability to identify specific strengths and weaknesses of the system. This limitation was addressed through the addition of patient- and clinician-specific feedback surveys that inquired about specific aspects of the system. Clinicians’ favorable ratings on items pertaining to the ease of locating sensor data in Epic and using the SmartPhrase in their clinical documentation aligns with their frequent viewing of sensor data. Patients’ mixed responses on the ease of data syncing aligns with the median daily adherence of 69% and were likely influenced by the transition from automated to manual syncing, a more burdensome process for patients. Although clinicians reported that sensor data added value to their patient care-corroborated by a 42-day median monitoring duration-patients’ perceptions of its personal usefulness were mixed. This was likely because, when queried further, many patients reported that they did not prefer using the clinician note in MyChart to view their sensor data due to the additional steps involved or forgetting data were there. Together, these findings highlight the importance of tailoring digital health solutions to end-user preferences. Integrating sensor data into clinicians’ preferred location (the EHR) yielded high data viewing and positive perceived utility. Conversely, data placement in the clinician’s note in MyChart was suboptimal for patients, which likely contributed to the variability seen in daily syncing adherence and lower perceived usefulness.

Considerable resources were required to build and maintain the wearable-to-EHR solution. In particular, research staff were needed to monitor syncing adherence, manually send reminders to sync, and provide tech support to patients and clinicians. Our finding that the majority (73.7%) of patients required at least one reminder to sync their data aligns with studies in rehabilitation^65^ and other populations^66, 67^, as does the median number of reminders (2) per patient.^65^ The digital health literature has emphasized the importance of providing adequate technical support to end-users^67–69^-a point supported by our data, where the majority of clinicians (15/17) and many patients (8/38) required technical assistance from the research team. The resource burden experienced in this study could be minimized in future work through selection of a less costly sensor, establishing institutional protocols for projects requiring EHR integration to minimize build times, and leveraging automated approaches for data syncing, sending syncing reminders, and providing basic tech support. As the field of rehabilitation modernizes through digital health, budgeting for adequate time, funding, personnel, and technical support will be critical for successful implementation and sustainability.

This study provides critical data that will inform the development and testing of remote therapeutic monitoring (RTM) programs in rehabilitation.^70, 71^ Studies in the cardiovascular, endocrinology, and oncology fields have shown promise for improved patient management and reduced hospital readmissions with remote monitoring programs.^72–75^ For the rehabilitation field to realize these potential benefits, assessing feasibility, usability, and resource demands is critical to guide future scaling efforts. RTM programs that rely on EHR integration will not be worth the resources to make scalable if patients and clinicians do not engage in their necessary actions (e.g., data syncing by patients, data viewing by clinicians). Questions related to the utility of RTM data in practice will best be answered with usable systems that allow for recent data to be available prior to clinic visits and in a seeable location to promote data viewing. The fact that clinicians recruited patients with a wide range of diagnoses (Table 2) suggests that investing in these considerations is worthwhile as it highlights the potential of RTM programs to be useful for patients with a wide variety of conditions and across clinics who serve different patient populations. Although appropriate for proof-of-concept testing, the wearable-to-EHR solution used in this study was built around a research-grade sensor that is inaccessible to consumers. As RTM research evolves from small, proof-of-concept to larger-scale implementation studies, sensor selection should prioritize scalability by leveraging consumer devices accessible to patients.

### Limitations

There are several limitations of this study. Although the user-centered design approach yielded a digital health solution tailored to clinician and patient preferences, technical constraints precluded our ability to incorporate all feedback. The research-grade LEAP sensor does not permit the wearer to view data on the device itself or on the mobile app. We attempted to overcome this limitation by providing patients with the option to view their sensor data within the clinician note in MyChart. We learned, however, that patients did not prefer this option, which may have negatively affected syncing adherence and usability ratings. Providing patients with feedback in a more preferrable location would likely improve adherence to data syncing and usability ratings. Relatedly, incomplete data transfers that occurred with automated syncing necessitated a transition to manual syncing, which increased patient burden and likely negatively impacted syncing adherence and usability ratings. Minimizing patient burden through automated syncing is preferrable.^76, 77^ Although the sample size was intentionally small due to the proof-of-concept nature of this study, this also means that findings from this study may or may not generalize to other clinical environments. The wearable-to-EHR solution was built around the preferences of clinicians and patients within a single academic medical center where buy-in and perceived importance of measuring activity outside the clinic was high (bottom row(s) of Tables 1 and 2). Findings from this study therefore likely represent a more optimistic perspective on the feasibility and usability of wearable-to-EHR digital health solutions in rehabilitation clinical care.

## CONCLUSIONS

By combining user-centered design principles with dedicated resources, we built a feasible and usable wearable-to-EHR digital health solution to bridge the gap between brief clinic visits and daily life in outpatient rehabilitation care. Future work is needed to refine digital health solutions that rely on wearable-to-EHR integration. These refinements may include providing patients with more frequent feedback from the sensor and in their preferred viewing location as well as automated processes for data syncing and reminders to minimize patient and staff burden. Next steps include improving visualization of sensor data in the EHR and the creation of documentation templates to standardize how and where data are documented in the clinical note to facilitate analyses of how data are being used in practice. Limitations in scalability could be addressed through a bring-your-own-device model that leverages patients’ personal devices, allowing them to continue to use their device beyond the episode of care.^78^ Ultimately, these data and future refinements will allow the field to determine the impact of remote therapeutic monitoring programs on clinical decision-making and patient outcomes.

## Acknowledgements

We are extremely grateful to the patients and WashU clinicians (Theresa Notestine, Elizabeth Hughes, Joan Scacciaferro, Gretchen Vandercar, Seung Yeon Son, Hannah Schweickart, Deborah Priluck, Beth Crowner, Amy DeFranco, Kathleen Dolan, Kayley Stock, Jeffrey Lanter, Sarah Schuman, Rachel Logan, Kate Mueth, Taylor Barry, and Caroline Canova Hew) who participated in this project.

## Author Contributions

AEM- conceptualization, data curation, formal analysis, funding acquisition, investigation, methodology, project administration, visualization, writing- original draft, writing- review & editing

CLH- conceptualization, investigation, writing- original draft, writing- review & editing

MDB- conceptualization, investigation, writing- review & editing

MMC- investigation, writing- review & editing

BJJ- formal analysis, writing- review & editing

KRL- methodology, writing- review & editing

EEFC- writing- review & editing

CAN- writing- review & editing

TMM- methodology, writing- review & editing

CEL- conceptualization, funding acquisition, methodology, resources, supervision, writing- original draft, writing- review & editing

## Statements and Declarations

### Ethical Considerations

The Washington University Human Research Protection Office approved the study.

### Consent to Participate

All participants provided written informed consent to participate in the study.

### Consent for Publication

Not applicable

### Data Availability

The data generated during the current study are available from the corresponding author on request.

---

**Supplemental Figure 1.**
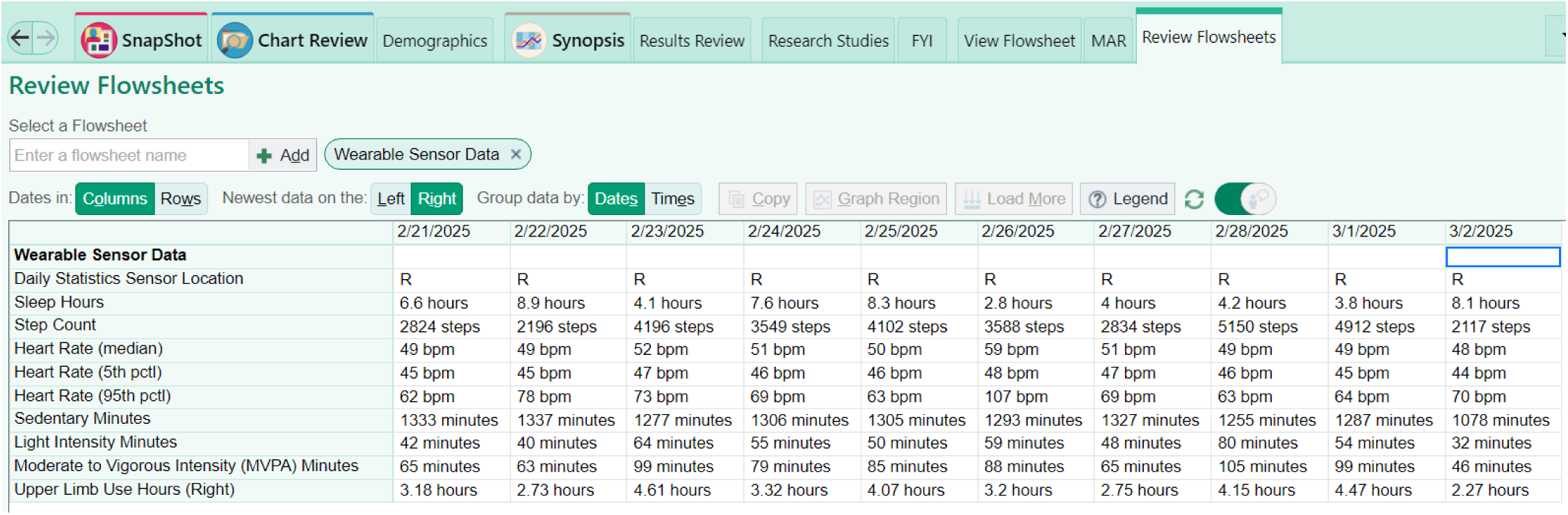
Clinician view of sensor data in the Epic flowsheet.

